# Effects of Improved Partner Notification on the Transmission of HIV and N. Gonorrhoea Among Men Who Have Sex With Men: A Modelling Study

**DOI:** 10.1101/2024.08.28.24312692

**Authors:** Maarten Reitsema, Jacco Wallinga, Birgit van Benthem, Eline Op de Coul, Ard van Sighem, Maarten Schim van der Loeff, Maria Xiridou

## Abstract

Men who have sex with men (MSM) are disproportionally affected by HIV in the Netherlands. Partner Notification (PN) is an important element of controlling the transmission of sexually transmitted infections (STIs) and HIV. We investigated the effects of improving PN on the transmission of HIV and N. gonorrhoeae (NG) among MSM in the Netherlands.

We developed an agent-based model that describes the transmission of HIV and NG among MSM. In the baseline scenario 14.3% and 29.8% of casual and steady partners of the index case get notified and tested for HIV/STI after three weeks (percentage notified and tested, PNT). We examined the following scenarios: 1) Increase PNT to 41% for both partner types; 2) Decrease the time between index and the partners tested to one week; 3) Combine scenario 1 and 2. Effects are expressed as cumulative change from the baseline simulation over 15 years.

Increasing PNT could lead to a decrease in gonorrhea cases of 45% (IQR: 39.9 – 49.9%), with an increase in number of HIV/STI tests of 4.4% (IQR: 1.6 – 7.3%), but no change in HIV infections (−5.4%; IQR: -21 – 7.9%). Decreasing time between tests could lead to a change in new NG infections of -14.2% (IQR: -17.2 – -10%), no change in HIV infections (8.2%; IQR: - 1.3 – 20%) or in number of HIV/STI tests performed (−0.4%; IQR: - 1.5 – 0.6%). Scenario 3 led to a change in NG infections of -56.8% (IQR: - 63.8 – - 47.4%), no change in HIV infections (11.5%; IQR: -11.1 – 33.9%) or in number of HIV/STI tests (− 0.5%; IQR: - 4.9 – 4.3%).

Increasing the percentage of sexual partners notified and tested for HIV/STI may have only a small effect on HIV, but could reduce the number of new NG infections substantially. However, it could lead to an increase in the number of HIV/STI tests performed.

**Key messages:** What is already known on this topic: Notifying recent sexual partners of people diagnosed with HIV or other sexually transmitted infections (STI) can promote timely testing and early treatment, thereby reducing further transmission of STIs

What this study adds: We quantify the effects of improving partner notification on the transmission of two STIs simultaneously.

How this study might affect research, practice or policy: In our scenarios, increasing the percentage of partners who get notified and tested had a bigger impact than decreasing the time between testing the index and his partners.

## Introduction

The annual number of HIV diagnoses in men who have sex with men (MSM) in the Netherlands has been decreasing since 2010. Overall, the total number of annual new diagnoses in the Netherlands has been less than 500 since 2020; and 86% of the total estimated population with HIV and 92% of those diagnosed and linked to care had a suppressed viral load (1). MSM are still disproportionately affected by HIV infections, with MSM accounting for 54% of HIV diagnoses in 2022 (1). The decrease in HIV diagnoses is in part the result of several policy changes that were made to curb the transmission of HIV. Pre-exposure prophylaxis (PrEP), has been shown to reduce HIV incidence by 86% (2, 3). In the Netherlands, PrEP can be prescribed since 2016, but has also been available for high risk MSM via a research project starting in 2015 in Amsterdam (4). A nationwide PrEP pilot was implemented in 2019, providing PrEP medication and care to individuals at high risk for HIV infection (5). Since 2015, it is recommended that HIV treatment is initiated as soon as possible after diagnosis. Early initiation of treatment is beneficial for the treated individuals (6, 7), and also for reducing onwards transmission (8). However, early diagnosis of individuals with HIV is needed for early initiation of treatment. Via partner notification (PN), sexual partners of newly diagnosed individuals (‘index’) are informed of their potential exposure to HIV and they are invited to services for testing. Notifying recent sexual partners of people diagnosed with HIV or other sexually transmitted infections (STI) can promote timely testing and early treatment, thereby reducing further transmission of STIs (9). Thus, PN is an important element in the work of STI clinic professionals. The aim of this study was to estimate the effects of improving PN on the spread of HIV and other STI using a mathematical transmission model. We included infections with Neisseria gonorrhoeae (NG) as an example of an STI other than HIV, as that is the most frequent STI among MSM in the Netherlands (5).

## Materials and Methods

### The transmission model

We updated an earlier published stochastic agent-based model (10-12) to simulate the spread of HIV and anogenital NG within the sexual network of MSM in the Netherlands. In short, the model simulates MSM 15-64 years old who have casual and steady sexual relationships with other MSM dependent on their respective ages, (known) HIV serostatus, and level of risk behavior. Within a sexual relationship, men can have sex with or without a condom. Through condomless anal intercourse MSM may acquire HIV and/or NG. After acquiring NG, the infection can be either symptomatic or asymptomatic. An infection with HIV starts with an acute phase of one month with a relatively high transmission rate, followed by a chronic phase. MSM get STI/HIV tested with varying intervals depending on their known HIV-status and sexual activity. Once diagnosed with HIV, an MSM enroll in HIV care and receive antiretroviral therapy (ART), which in time results in viral suppression and reduces the transmissibility of HIV to zero. The model is described in depth in the supplement.

### Model calibration

The model was calibrated to the annual number of new HIV diagnoses and annual positivity rate of anogenital gonorrhea for the years 2017-2019 (13-16). Using Latin Hypercube Sampling, we sampled 5,000 parameter sets from broad uniform distributions for eight free parameters (See supplement). Each parameter combination was run 20 times. Next, we calculated for each parameter set a relative distance 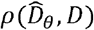, between observed values *D* and model outcomes 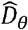 for annual number of HIV diagnoses and annual positivity rate of anogenital gonorrhea as 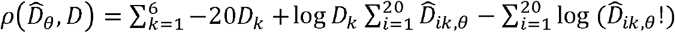, with *k* being the 6 data points to which the model is fitted, and *i* being the 20 replicate runs for each parameter set θ. This function is the log-likelihood function of the Poisson distribution, but used as a distance measure in an approximate Bayesian computation because the spread of two different diseases in different units are fitted concurrently. The 100 parameter sets that yielded the smallest distances were selected (Figure 1). For all scenarios, each parameter set was run 20 times for stochasticity.

**Figure 1.**
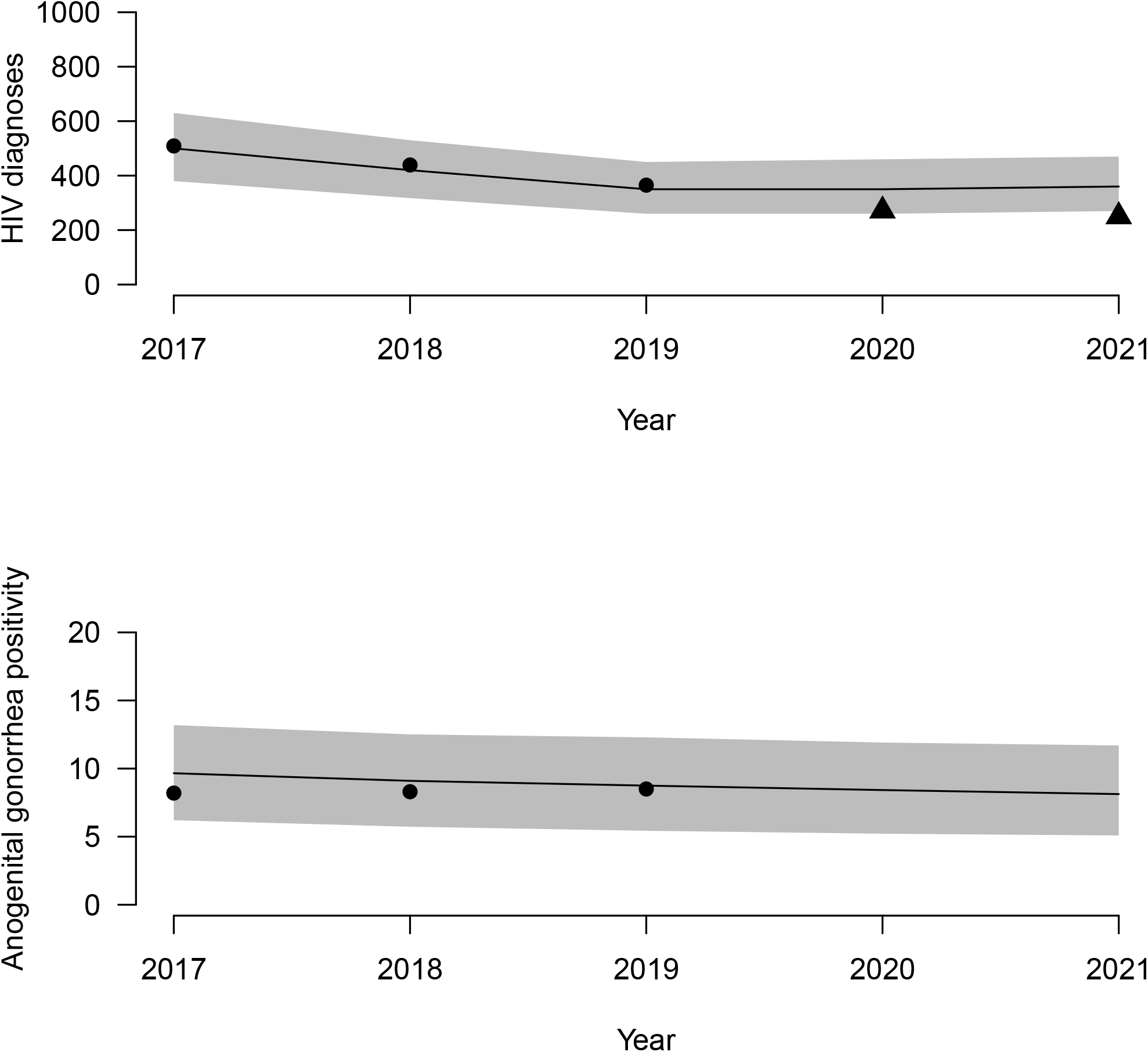
Model calibration and validation, based on the annual HIV diagnoses (top) and the positivity rate of anogenital gonorrhea (bottom). Lines and gray area: median and interquartile ranges calculated from the model. Markers show respective results from data (13-16), used for calibration (circles) or validation (triangles).

### Current Partner Notification

Within the model, once an individual is tested positive for HIV or NG, PN takes place. Casual and steady partners of the index have 14.3% and 29.8% chance, respectively, of getting notified and coming in for an HIV/STI test (percentage notified and tested: PNT) (18). The index’s partners will be tested for both HIV and NG regardless of the index’s diagnosis, unless the partner had already been diagnosed with HIV. In concordance with Dutch guidelines, only sexual partners in the preceding 5 weeks or 6 months are eligible to be notified following a diagnosis of symptomatic or asymptomatic gonorrhea, respectively (19). For HIV, guidelines suggest contacting partners as far back in time as possible, but for computational considerations we limited this to partners in the last two years. For the years 2020 – 2035, we examined the following changes in partner notification (referred to as interventions):

1. Increasing the PNT: The PNT is increased for both casual and steady partners to 41%. This is based on the practical upper limit of the percentage of partners that can be notified and tested, as some partners are anonymous or in some other way not notifiable (18).
2. Decreasing the time between testing the index and his partners from 3 weeks to 1 week.
3. Combining scenario 1 and 2: PNT was increased to 41% for both steady partners and casual partners, and the time between the test of the index and his notified partners was reduced to 1 week.

In sensitivity analyses, we examined the impact of our main scenarios in the following way:

1a. Increasing the PNT to 41%, only for steady partners
1b. Increasing the PNT to 41%, only for casual partners
1c. Increasing the PNT to 41%, only after an HIV diagnosis.
1d. Increasing the PNT to 41%, and increasing the time between tests to 4 weeks. 2a. Decreasing the time between tests to 2 weeks.

We calculated the cumulative number of new HIV infections, NG infections, and HIV/STI tests performed during the first 15 years from the moment the intervention was introduced. As outcome measure, we calculated the percentage change in these numbers compared to simulations without intervention (baseline scenario). We present results as the median and interquartile range (IQR); the 95% credibility interval are shown in the supplement. A reduction or negative value indicates fewer infections or HIV/STI tests in the scenario of interest compared to the baseline scenario.

## Results

### Scenario 1, increasing percentage notified and tested

An increase in the PNT led to a 45% (IQR: 38.9 – 49.9%) decrease in the cumulative number of new NG infections (Figure 2). However, in the same period, there was an increase in the numbers of STI tests performed of 4.4% (IQR: -1.6 – 7.3%), but no change in HIV infections (−5.4% IQR: -21 – 7.8%). The number of tests performed increased sharply in the first year of the intervention (6.6%; IQR: 5.6 – 7.6%), as more partners of MSM diagnosed with an HIV/STI were notified and tested. This effect decreased over time as the intervention concurrently led to fewer new NG infections: the change in new NG infections in 2020 was -17% (IQR: -19.7 – -13.4%).

**Figure 2.**
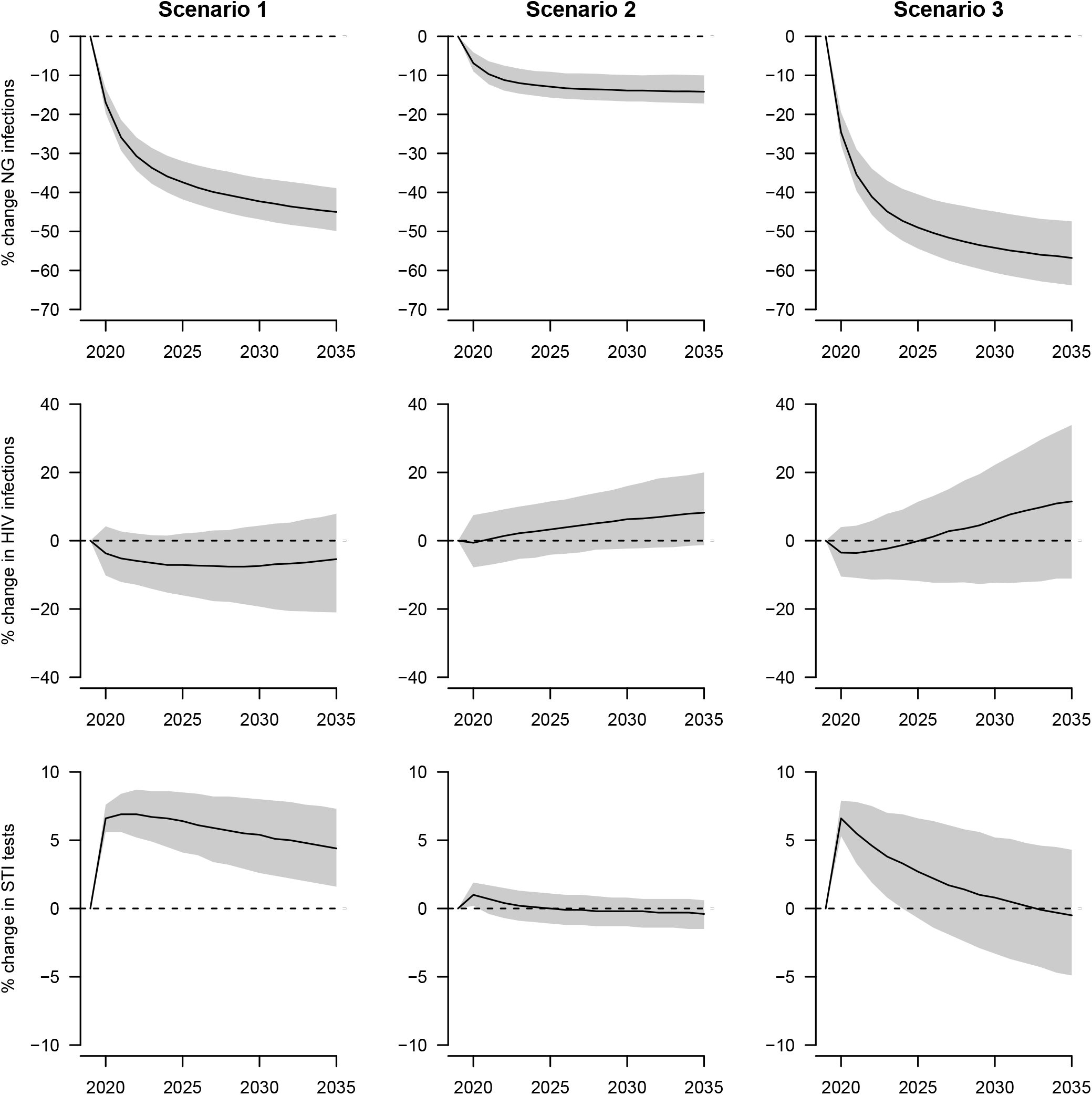
Effect of changes in partner notification. The left column shows scenario 1 (increased percentage of partners notified and tested (PNT)); the middle column shows scenario 2 (decreased time between testing index and partners); the right column shows scenario 3 (combination of 1 and 2: increased PNT and decreased time between testing index and his partners). The top row shows the percentage change in cumulative number of new NG infections; the middle row shows the percentage change in cumulative number of new HIV infections; the bottom row shows the percentage change in cumulative number of new HIV/STI tests performed. The solid line and grey area show the medians and interquartile ranges. The dotted lines denote no change (0% change in scenario 1, 2, or 3, compared to the baseline scenario).

### Scenario 2, reducing time between testing index and partners

Reducing the time between testing the index and testing the index’s partners resulted in a decrease in number of new NG infections, without a change in HIV/STI tests performed or any change in new HIV infections. The cumulative number of NG infections changed by -14.2% (IQR: -17.2 – -10.0%) over a 15 year time period, which was less than scenario 1. In the same period, there was hardly any change in STI tests performed (−0.4%; IQR: -1.5 – 0.6%), or in HIV infections (8.2%; IQR: -1.3 – 20%).

### Scenario 3, Increasing PNT and reducing time between index and partners

Increasing the success rate and decreasing the time between tests could lead to a reduction in new NG infections of 56.8% (IQR: 47.4 – 63.8%) over a 15 year time period. This reduction was larger than the respective reductions in scenario 1 and 2. There was an increase in HIV/STI tests in the first year of 6.6% (IQR: 5.3 – 7.9%), but this effect disappeared over time. At the end of the simulated 15 years, there was no change in STI tests performed (−0.5%; IQR: -4.9 – 4.3%). Additionally, there was no change in HIV infections (11.5%; IQR: -11.1 – 33.9%).

### Sensitivity analysis

When applying the increased PNT (scenario 1) only to steady partners of the index case, there were no changes in the cumulative number of HIV/STI tests performed, nor in the cumulative number of new HIV and NG infections compared to the baseline scenario (Table 1). However, when applying the same scenario only for casual partners of the index patient, the changes over 15 years are similar to the results in scenario 1, with a change in NG infections of -44.7% (IQR: -49.9 – -38.2%). When we assumed that the increase in PNT would require an extra week between testing the index patient and their partners, there would be a decrease in number of new NG infections (− 37.8% (IQR: - 42.6 – -32.8%)) and increase in number of HIV/STI tests (6.1% (IQR: 3.6 – 8.3%)). Interestingly, the increase in time between tests could lead to a larger median percentage decrease in HIV infections.

**Table 1.** Results of the sensitivity analyses for the impact of improvements in partner notification on the numbers of HIV and NG infections and the numbers of STI/HIV tests.

|  | STI/HIV tests <sup>a</sup> | HIV infections <sup>b</sup> | NG infections <sup>c</sup> |
| --- | --- | --- | --- |
| **Scenario 1 <sup>\*</sup>** |  |  |  |
| *Main Scenario* | 4.4% (IQR: 1.6 – 7.3%) | -5.4% (IQR: -21 – 7.9%) | -45% (IQR: -49.9 – -38.9%) |
| *Only steady* | 0.3% (IQR: -0.4 – 1.1%) | -1.7% (IQR: -10.5 – 7.7%) | -0.9% (IQR: -3 – 1.3%) |
| *Only casual* | 4.4% (IQR: 1.7 – 7.2%) | -3.8% (IQR: -19.9 – 11.4%) | -44.7% (IQR: -49.4 – -38.2%) |
| *Only HIV* | 0.5% (IQR: -0.3 – 1.5%) | -6.2% (IQR: -14.5 – 4.3%) | -1.4% (IQR: -3.7 – 0.9%) |
| *4 weeks* | 6.1% (IQR: 3.6 – 8.3%) | -11.9% (IQR: -24.7 – 0.8%) | -37.8% (IQR: -42.6 – -32.8%) |
| **Scenario 2 <sup>\*\*</sup>** |  |  |  |
| *Main Scenario* | -0.4% (IQR: -1.5 – 0.6%) | 8.2% (IQR: -1.3 – 20%) | -14.2% (IQR: -17.2 – -10%) |
| *2 weeks* | 0.0% (IQR: -0.8 – 0.8%) | 3.4% (IQR: -5.6 – 14.2%) | -6.6% (IQR: -9 – -3.9%) |

<sup>a</sup> percentage change in cumulative number of STI/HIV tests performed over 15 years.
<sup>b</sup> percentage change in cumulative number of HIV infections over 15 years.
<sup>c</sup> percentage change in cumulative number of NG infections over 15 years.
<sup>\*</sup> **Only steady:** Scenario 1, but only applied to steady partners of the index patient; **Only casual:**
Scenario 1, but only applied to casual partners of the index patient; **Only HIV:** Scenario 1, but only
applied after an HIV diagnosis in the index patient, and not after a gonorrhoea diagnosis. **4 weeks:**
Scenario 1, but with an increased time between testing the index patient and their partners from 3 weeks to 4 weeks.
<sup>\*\*</sup> **2 weeks:** Scenario 2, but time between the test of the index and their partners is reduced from three to two weeks instead of to one week.

The results of scenario 2 were attenuated if the time between the test of the index and the test of their partners was decreased to 2 weeks instead of 1 week.

## Discussion

In this study, we modelled two forms of improvements to PN: increasing the percentage of partners that get notified and tested, as well as decreasing the time between the test of the index and his partners. Our results showed that implementing separately one of these improvements could result in a reduction in the number of new NG infections. Increasing the PNT could lead to an increase in the number of HIV/STI tests performed, although this increase in HIV/STI tests diminished over time, possibly due to the reduced number of new NG infections over time. More NG infections could be averted if the increase in PNT was combined with a decrease in time between tests, and in this scenario, the elevation in HIV/STI tests due to the intervention disappeared within the simulated 15 years.

According to our results, increasing the PNT could lead to a bigger decrease in new NG infections than decreasing time between tests. In our model, partners of the index were randomly selected to get tested. When the PNT is increased, the partner who transmitted an STI to the index is more likely to present for an HIV/STI test. Additionally, by increasing the PNT, more MSM were tested which can lead to opportunistically finding more infections. Finally, with the higher probability of finding cases, again more effective PN is initiated. Conversely, by decreasing the time between tests, the infectious period of a partner could be shortened, but only if the partners notified and tested are the ones with HIV/STI.

Our simulations showed little effect of changes in PN on the number of new HIV infections. In fact, an upwards trend was noticeable when looking at the medians of scenarios 2 and 3. The upward trend could be caused by testing partners too soon after transmission, when a test would yield a false negative result. The sensitivity analyses appeared to corroborate this, as this upward trend was attenuated when the time between tests was decreased from 3 weeks to 2 weeks instead of to 1 week (sensitivity analysis 2a), and by the more pronounced effect of increased PNT on HIV transmission if an extra week was added between tests (sensitivity analysis 1d). This is a limitation of our study, as health care workers would know to wait for the appropriate time after a potential exposure event to HIV for a test. The overall small effects on HIV transmission compared to the effects on NG transmission could be caused by several factors. First, men who are diagnosed with gonorrhea were infected within the period from which (ex)-partners are selected: 5 weeks for symptomatic gonorrhea and 6 months for asymptomatic gonorrhea. On the other hand, after an HIV diagnosis only (ex)-partners in the last two years were selected for PN, while the index could have acquired HIV from, or could have transmitted to, a sexual partner before that time period. This is a limitation of our model and could lead to an underestimation of the effectiveness of PN on HIV. Second, the transmission rate and thus the incidence of HIV is lower than the incidence of gonorrhea. A study has found that gonorrhea incidence was positively correlated with the percentage of partners notified and tested that had an NG infection (20). Yet, PN still has a role to play in HIV prevention as it might link men to care who would not get tested on their own. Additionally, according to data of Sexual Health Centres in the Netherlands in 2023, the HIV positivity rate was higher for MSM who had been notified for HIV (2.0%) than those who had not been notified (0.2%) (21).

We have made several assumptions in the model. First, we modelled the PNT as independent of the time since a relationship ended in both the baseline and intervention scenarios. Intuitively, more recent partners may be notified more reliably than partners with whom the relationship ended longer ago. If the PNT was higher for more recent partnerships, the effect on NG might be higher than presented here, as the transmission event of NG is at most 6 months ago. Moreover, The ExGen study on the natural history of rectal NG infections found median a duration of 9 weeks, based on 20 infections of which 14 were censored due to treatment (22), which also shows the methodological difficulties estimating the natural duration of gonorrhea. If the natural duration of gonorrhea is considerably shorter, and only partners from a shorter time window need to be notified, the effect of increased PNT on NG could have been underestimated in this study. Still, current guidelines for PN for asymptomatic gonorrhea advice notifying partners of the last 6 months.

Our model does not include antibiotic resistance. NG has been shown to be capable of developing antimicrobial resistance (23). Resistance to ceftriaxone, the first line antibiotic, has been observed in various countries (24), but not yet in the Netherlands (25). Increasing levels of antimicrobial resistance of NG over time could mean that we overestimate the effect of PN on gonorrhea transmission if the development of resistance would lead to treatment failures.

Simulating the effects of PN using an agent-based model with two STIs allowed us to capture the non-linear impact of PN. Our method ensured only individuals that had a sexual interaction with the index were eligible for HIV/STI testing, and allowed for opportunistic diagnoses to take place, even of a STI different from the index case. These dynamics cannot be captured in compartmental models, which typically add to a diagnosis rate to estimate the effects of PN (26, 27), which could possibly overestimate the effect on HIV/STI.

In conclusion, increasing the PNT and/or decreasing the time between tests had little effect on HIV transmission in our model, but the interventions could reduce the number of new NG infections substantially. Increasing the PNT could lead to an increase in the number of HIV/STI tests performed.

## Data Availability

All data produced in the present study are available upon reasonable request to the authors

## Funding

This work was supported by Aidsfonds grant number 2014037

## Contributors

MR, JW, BvB, EodC, AvS, MSvdL and MX conceived and planned the study.

MR programmed the mathematical model and carried out the modelling

MR, JW, BvB, EopC, AvS, MSvdL, MX contributed to the interpretation of the results.

MR took the lead in writing the manuscript. All authors provided critical feedback and helped shape the research, analysis and manuscript.

MR is the guarantor.

## Competing Interests

None declared

## Ethics Approval

Not applicable / No human participants included

## Notes

### Competing Interest Statement

The authors have declared no competing interest.

