## Supplement for "Effects of Improved Partner Notification on the Transmission of HIV and N. Gonorrhoea Among Men Who Have Sex With Men: A Modelling Study"

**Contents**

| The transmission model | 3 |
| --- | --- |
| Demographics | 3 |
| Steady partnerships | 3 |
| Casual partnerships | 4 |
| Mixing according to HIV status: serosorting | 4 |
| Mixing according to age and risk group | 5 |
| Condom use | 5 |
| HIV infection | 5 |
| Gonorrhoea | 5 |
| HIV/STI testing | 5 |
| Partner notification | 6 |
| Calibration | 6 |
| Figure S1: Parameter values sampled and selected via calibration | 7 |
| Figure S2: Results with 95% credibility interval. | 9 |
| Tables S1-S5: Values, distributions, and sources for model parameters | 10 |
| References | 15 |

**The transmission model**

Our model describes the transmission of HIV and *Neisseria gonorrhoeae* (NG) among MSM. For brevity, we use the terms HIV and gonorrhoea to refer to infection with the pathogen and to specific diseases caused by the pathogen. In this model, we follow MSM, who can have steady and/or casual partners and can have condomless anal intercourse (CAI) or other types of sex acts. We assumed that HIV and gonorrhoea can only be transmitted via CAI. The model integrated data from several data sources, including the Amsterdam Cohort Study (ACS) among MSM (1), the Network Study among MSM in Amsterdam (2), the national database of HIV positive individuals in the Netherlands (3), the national database of STI clinics in the Netherlands (4). The time step in the model was one day. The model has been written in JAVA using Eclipse NEON, and run using openjdk version 1.8.0_152-release. For statistical distributions the commons-math3 3.6.1 package was used. The runtime of one scenario with 2,000 stochastic realizations is around 130 hours when not run in parallel.

**Demographics**

The model population consists of 20,000 individuals. Model results were scaled to the estimated MSM population in the Netherlands of 200,000 (5). We simulate MSM aged 15-64 years old, with ages uniformly distributed. A uniform distribution was chosen as the age distribution of MSM in the Netherlands can be expected to be different from the general population, as AIDS had a significant impact on premature mortality in 1982-1996 (6), and self-acceptance of homosexual urges might be different for different age groups. Individuals enter the MSM population when they are 15 years old, but they may not be sexually active at that time. The distribution of age at sexual debut was estimated from ACS data using maximum likelihood techniques (**Table S1**). An individual leaves the sexually active MSM population upon reaching the age of 65. We assumed a constant population size so that the number of individuals who enter the population equals the number of individuals leaving the population.

**Steady partnerships**

After sexual debut, MSM in the model can have up to one steady partner on any given time point. We estimated that 65% of MSM had a steady partner in the preceding six months based on data from the ACS and the Network Study. When the percentage of MSM who had a steady partner in the preceding six months is lower than 65%, new steady partnerships are formed. Potential partners are recruited from the MSM population in a random order; the rate of recruitment depends on the number of MSM with and without steady partner and an assumed proportionality factor that makes some MSM more likely to start steady partnerships than other MSM. This factor is equal to 0.5 for 40% of MSM, 1.0 for 50% of MSM, and 2.0 for 10% of MSM. This means that 10% of MSM were twice more likely and 40% of MSM are 50% less likely to start a steady partnership than the remaining 50% of MSM. The duration of steady relationships and the frequency of sex acts within steady relationships were sampled from distributions fitted to data from ACS and the Network Study respectively, using maximum likelihood estimation (**Table S1**). When the relationship is formed, a value for its duration is sampled from the relative distribution. The relationship ends when this duration is reached or when one of the two agents reaches the age of 65 or develops AIDS. A sex act rate is drawn from a beta distribution fitted to data from the Network Study. (**Table S1**).

**Casual partnerships**

After sexual debut, MSM in the model can have casual partnerships, even on days that they have a steady partner. For every individual, we define the “sexual desire” as the number of casual partners that he would like to have in six months. Men who had fewer partners in the last six months than their sexual desire are recruited for new casual relationships. Relationships are not terminated prematurely when the sexual desire decreases. The sexual desire is sampled from one of five distributions (**Table S1**); where the five distributions differ for MSM in the five age groups (15-24, 25-34, 35-44, 45-54, 55-64). The distributions of the age groups 15-24 and 25-34 were inferred from data from the ACS. The distributions for the three older age groups 35-44, 45-54, 55-64 were based on the distribution of 25-34 year-olds, with the mean parameter shifted by a factor determined via the calibration process, to ensure that the distribution of HIV diagnoses according to age matches the respective distribution from SHM data. The number of sex acts and the duration of casual relationships were inferred from data from the Network Study (**Table S1**). A casual relationship always starts with a sex act, to ensure at least one sex act per casual relationship. The median length of a casual relationship is 1 day, while the median number of acts per day, excluding the first act, is 0.056. The probability that a man will not use a condom is sampled from a distribution fitted to data from the ACS (**Table S1)**. When two men have intercourse, the lowest probability for condomless sex is used.

**Mixing according to HIV status: serosorting**

Parameters for serosorting were based on data from the Network Study. From these data we calculated that among diagnosed HIV-positive MSM who start a steady relationship, 53.5% forms this relationship with another diagnosed HIV-positive MSM and the remaining 46.5% with another MSM randomly chosen. MSM who have had a negative HIV test in the preceding year or who have been sexually active for less than a year have a chance of 63.9% of starting a steady partnership with a man with the same characteristics; however, these individuals can have acquired HIV if the infection took place more recently than the HIV test. For casual partnerships, the percentage serosorting was 26.1% for diagnosed HIV-positive MSM and 14.2% for MSM who had a negative HIV test in the last year (**Table S2**).

**Mixing according to age and risk group**

When a steady or casual partnership is formed in the model, the choice of a partner depends on the age of the men involved. Mixing according to age is based on data from the Network Study, adjusted to relative rates using maximum likelihood and taking reciprocity of sexual acts into account (7) (**Table S3)**, as follows. Let *m_jk_* denote the fraction of men of age-group *k* that forms partnerships with men of age-group *j*, that is the element in the *j*-th row and *k*-th column of the matrix shown in Table S3. Let *N_k_* denote the number of men in age-group *k*. Then *m_jk_* were calculated from the data such that *m_jk_ N_k_* = *m_kj_ N_j_* and the sum of the elements of each column equals 1, for all *j* and *k*. This ensures that the total number of partnerships formed by all men of age-group *j* with men of age-group *k* equals the total number of partnerships formed by all men of age-group *k* with men of age-group *j*. We define high-risk MSM as those having more than 20 partners in six months and low-risk as those with up to 20 partners in six months. Based on ACS data, approximately 10% of MSM is high risk. We assume a level of 75% of assortativeness: 75% of casual relationships of high-risk MSM is with high-risk MSM and 75% of casual relationships of low-risk MSM is with low-risk MSM. The remaining 25% of both groups are matched proportionally.

**HIV infection**

The probability, *p_H_(V)*, of HIV transmission per act of CAI with an HIV-positive man with viral load *V* log_10_(copies/mL) was calculated as $p_{H}(V)={2.45}^{V- 4.5} \beta_{H}$ (8, 9). In this formula, $\beta_{H}$ is the probability of HIV transmission when HIV viral load is 4.5 log­_10_(copies/ml). The value of $\beta_{H}$ is obtained via the calibration process. If the HIV-positive MSM is also infected with gonorrhoea, *p_H_(V)* is multiplied by a factor of 1.17 (8). The first 30 days after transmission, form the acute phase with high viral load (10). After that, HIV viral load declines, forming the chronic phase (**Table S1**). We assumed that ten years after acquiring HIV, untreated individuals develop AIDS. Due to the severity of AIDS, we assumed that individuals with AIDS do not engage in risky sexual practices and do not contribute to further transmission (11). Parameters relating to chronic HIV infection were fitted to data from SHM. When an individual is diagnosed with HIV, the time between diagnosis and viral suppression follows negative binomial distributions with a median time of 84 days. (**Table S4)**.

**Gonorrhoea**

We modelled only anogenital gonorrhoea. The probability of transmission of gonorrhoeae per act of CAI is $\beta_{G}$ and the probability of NG infection being symptomatic is *P_symptoms_*; the values of these parameters were obtained via the calibration. We assumed that MSM with symptomatic gonorrhoea always seek testing and care; they receive antibiotic treatment immediately when tested. MSM with asymptomatic gonorrhoea become infectious 8 days after infection; they may be tested opportunistically, but they receive treatment after test results are available. Undiagnosed gonorrhoea is cleared in 6 months (**Table S4)**.

**HIV/STI testing**

In two different surveys among MSM in the Netherlands (12, 13), approximately 20% of MSM reported that they had never been tested for HIV. Therefore, in the model, we assumed that 20% of MSM do not get tested, unless they have symptoms for gonorrhoea or they are notified by a partner. The remaining (80% of MSM) gets tested for HIV/STI once every 5 years, 2 years, or six months. These testing frequencies were determined based on data from the national database of STI Clinics in the Netherlands (14) (**Table S5)**. A new testing frequency is sampled for an individual if his sexual desire changes (i.e. when he ages to the following age group and/or when he starts/ends a steady partnership), or if he is diagnosed with HIV.

**Partner notification**

Partner notification is offered to MSM testing positive for HIV and/or gonorrhoea (15). These men are asked about their sexual partners and whether these partners can be notified and invited to get tested. In accordance with guidelines from the National Coordination Centre for Communicable Disease Control, only partners of the preceding six months are notified when MSM are diagnosed with asymptomatic gonorrhoea and partners of the preceding six weeks are notified when MSM are diagnosed with symptomatic gonorrhoea. Notified partners get tested after three weeks. Guidelines by Coordination Center for Communicable Disease Control provide no time limit for partner notification after an HIV diagnosis. However, in the model, we keep information about ex-partners for a maximum of two years after the end of a relationship, in order to keep up the computational speed of the model **(Table S2**).

**Calibration**

The model was calibrated to the annual numbers of HIV diagnoses and the positivity rate of anogenital gonorrhoea in 2017-2019 using an approximate Bayesian computation method. The positivity rate was defined as the percentage of gonorrhoea positive tests out of all gonorrhoea tests carried out (among MSM and excluding pharyngeal-only gonorrhoea). Using Latin Hypercube Sampling (16), we sampled 5,000 parameter combinations from broad uniform distributions for the eight free parameters:

1. HIV transmission probability (probability of acquiring HIV per sexual act)
2. NG transmission probability (probability of acquiring NG per sexual act),
3. NG symptomatic probability
   4-6. Parameters alpha1, alpha2, alpha3 shifting the distribution of sex partners of 25-34 year old MSM to obtain the distribution of partners of MSM 35-44, 45-54, 55-64 years old, respectively.
7. HIV import rates (probability of an MSM acquiring HIV from outside the Dutch MSM population, per day)
8. NG import rate, (probability of an MSM acquiring NG from outside the Dutch MSM population, per day)

The model calculations were repeated 20 times for each parameter combination to account for stochasticity. We calculated the distance ${\rho(\hat{D}}_{\theta},D)$, or level of discrepancy, between reported data $D$ and model output $\hat{D}_{\theta}$ (with parameters θ) as $\rho\left( \hat{D}_{\theta},D \right)=\sum_{k=1}^{6} -20D_{k}+\log D_{k}\sum_{i=1}^{20} \hat{D}_{ik,\theta}-\sum_{i=1}^{20} log(\hat{D}_{ik,\theta}!)$, with *i* denoting one of the 20 stochastic realizations and *k* denoting one of the 6 data points to which the model was calibrated (annual HIV diagnoses and anogenital positivity of gonorrhoea). This distance measure is log-likelihood function of the Poisson distribution. We selected parameters θ for which $\rho\left( \hat{D}_{\theta},D \right)<\epsilon$. We chose tolerance $\epsilon$ such that 100 out of the 5,000 parameter combinations were selected. Subsequently, model calculations were carried out with the 100 selected parameter combinations.
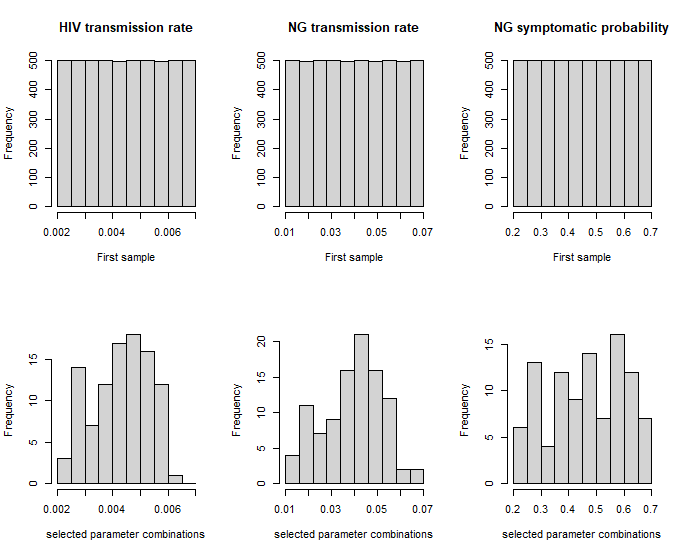

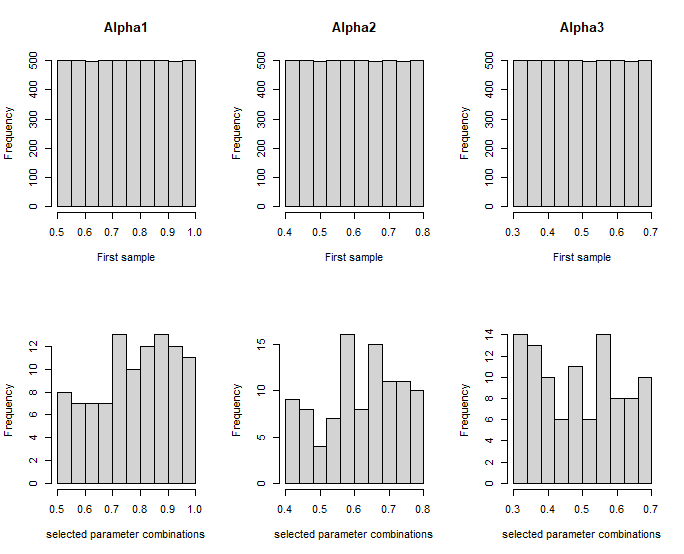

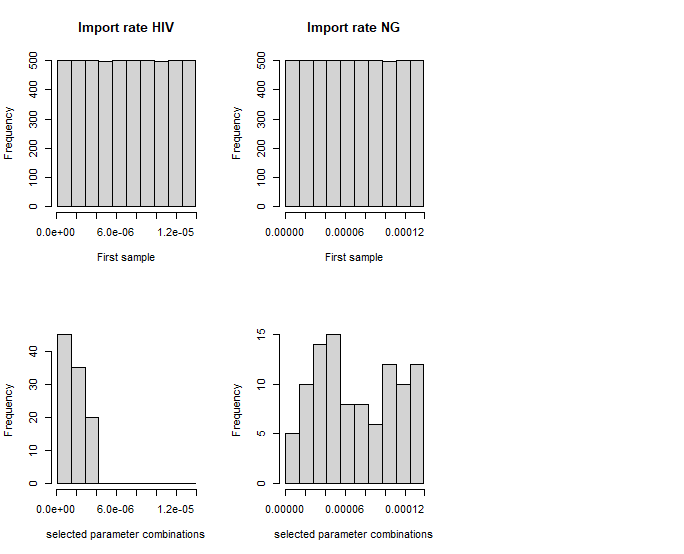

**Figure S1: Parameter values sampled and selected via calibration.** Parameters are: HIV transmission rate: probability of HIV transmission when log_10_ Viral load is 4.5; GO transmission rate: probability of NG transmission; GO symptomatic probability; alpha 1-3: scalars to adjust sexual activity of older age groups; HIV import rate: probability of MSM acquiring HIV not via the simulated sexual network; GO import rate: probability of MSM acquiring NG not via the simulated sexual network

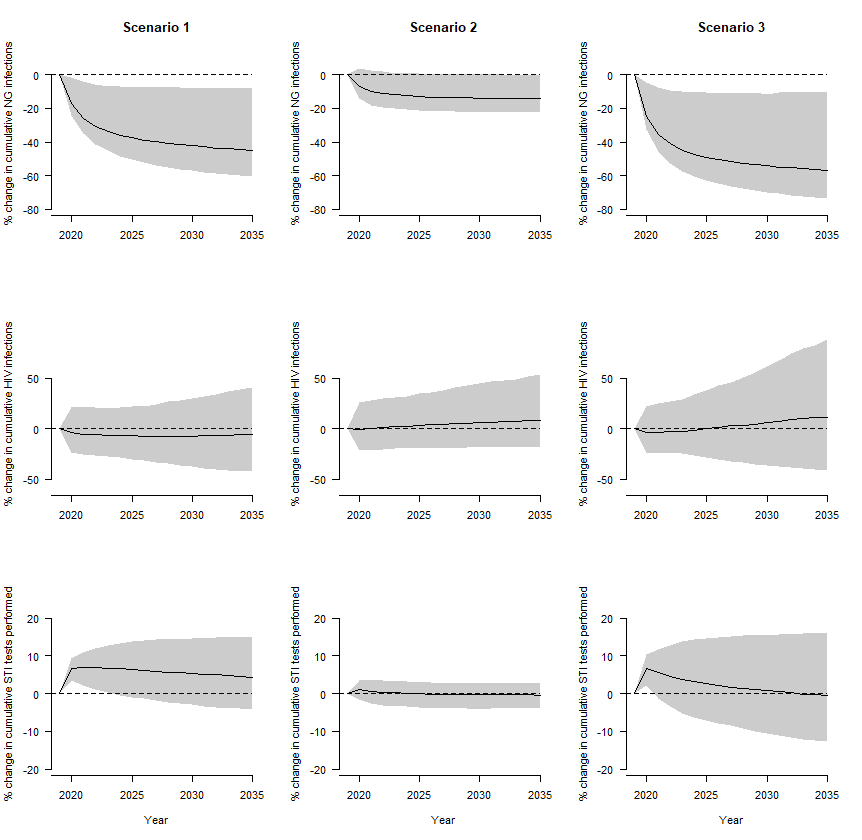

**Figure S2: Results with 95% credibility interval.** These results are identical to those shown in Figure 1 of the main text, but with the grey area showing the 95% credibility interval instead of the interquartile range. The 95% Credibility interval is defined as the interval in which 95% of the model simulations fall.

**Table S1.** Parameters of the transmission model with distributions.

| **Model parameter** | **Distribution^#^** | **Source^§^** | |
| --- | --- | --- | --- |
| *Steady partnerships* |  | |  |
| Length (days) | Weibull(shape = 0.60888, scale = 920) | | ACS |
| Number sex acts per day | Beta(α = 0.5615, β = 2.002) | | NWS |
| CAI chance | Beta(α = 0.1926, β = 0.2026) | |  |
| *Casual partnerships* |  | |  |
| Length (days) | Gamma (shape = 0.025758, rate = 0.000168) | | NWS |
| Number sex acts per day | Beta (α = 0.4805, β = 3.9098) | | NWS |
| CAI chance | Beta(α = 0.1926, β = 0.2026) | | ACS |
| *Number of casual partners* |  | | ACS |
| 15-24 years old | Nbinom(size = 0.364, mu = 9.394) | |  |
| 25-34 years old | Nbinom(size = 0.379, mu = 10.619) | |  |
| 35-44 years old | Nbinom(size = 0,538, mu = 14.474*alpha1^†^) | |  |
| 45-54 years old | Nbinom(size = 0,538, mu = 14.474*alpha2^†^) | |  |
| 55-64 years old | Nbinom(size = 0,538, mu = 14.474*alpha3^†^) | |  |
| *HIV RNA viral load (log_10_ copies/mL)* |  | |  |
| Acute HIV | 7.0 | | (10) |
| chronic HIV | Normal (mean = 4.5, sd = 0.7) | | SHM |
| Suppressed HIV | 0 | | SHM |
| *Time periods in HIV progression (days)* |  | |  |
| From diagnosis to suppression  Age at sexual debut (years) | Nbinom(size = 2.62562, mu = 96.42084)  Normal (mean = 17.5, sd = 3.4) | | SHM  ACS |

**^#^**Nbinom, Negative binomial distribution with parameters size and mean; Normal, normal distributions with parameters mean and standard deviation (sd); Beta distribution with parameters alpha and beta; Weibull distribution with parameters alpha and beta.

**^§^**Distributions obtained from data from the following studies: ACS, Amsterdam Cohort Study among MSM; NWS, Network Study among MSM in Amsterdam; SHM, Stichting HIV Monitoring.

† w1, w2 and w3 are scalers determined in the calibration process.

†† For suppression a relatively strict definition was used when analysing SHM data: 2 consecutive RNA measurements of less than 100 copies / ml.

cART, combination antiretroviral therapy; CAI, condomless anal intercourse.

**Table S2:** Parameters of the transmission model relating to sexual behaviour and partner notification.

| **Parameter** | **Value** | **Source** |
| --- | --- | --- |
| Serosorting with steady partners: |  |  |
| HIV-positive diagnosed | 53.5% | NWS |
| Negative HIV test in preceding 12 months or no sexual activity in preceding 12 months | 63.9% | NWS |
| Serosorting with casual partners: |  |  |
| HIV-positive diagnosed | 26.1% | NWS |
| Negative HIV test in preceding 12 months | 14.2% | NWS |
| Number of MSM in the Netherlands | 200,000 | (5) |
| Number of MSM in the model | 20,000 | * |
| Age of MSM accounted for in model | 15-64 years | * |
| Percentage of MSM having a steady partner in the preceding 6 months | 65% | ACS |
| % of high-risk** MSM choosing a high-risk man as casual partner | 75% | * |
| % of low-risk** MSM choosing a low-risk man as casual partner | 75% | * |
| Partner notification after HIV/STI diagnosis: |  |  |
| % of casual partners getting tested | 14.3% | (15) |
| % of steady partners getting tested | 29.8% | (15) |
| When an MSM is diagnosed with HIV or gonorrhoea, partners are notified if the relationship is still active, or if the relationship ended within this period: | | * |
| Asymptomatic gonorrhoea | 6 months |  |
| Symptomatic gonorrhoea | 6 weeks |  |
| HIV | 2 years |  |
| Time between partner notification and test of the notified partner | 3 weeks | * |

*Model assumption.

** High-risk MSM are those with more than 20 partners per six months; low-risk MSM are those with up to 20 partners per six months.

MSM, men who have sex with men; NWS, data from the Network Study among MSM in Amsterdam; ACS, data from the Amsterdam Cohort Study among MSM.

**Table S3.** Mixing of ages of partners in steady and casual relationships. The fractions shown in a column represent the fractions of partnerships formed by MSM of the age group indicated in that column with the age groups indicated in each row. The fractions were calculated using data from the Network Study on the number of partners of men of a specific age and the age of their partners. The data were adjusted to relative rates using maximum likelihood and taking reciprocity of sexual partnerships into account (see paragraph on mixing according to age).

| **Age of partners** | **Age of index MSM (years)** | | | | | | | | | | | | | | | | | | |
| --- | --- | --- | --- | --- | --- | --- | --- | --- | --- | --- | --- | --- | --- | --- | --- | --- | --- | --- | --- |
|  | 15-19 | | 20-24 | | 25-29 | | 30-34 | | 35-39 | | 40-44 | | 45-49 | | 50-54 | | 55-59 | | 60-64 |
| ***Steady partners*** | | |  | |  | |  | |  | |  | |  | |  | |  | |  |
| **15-19** | 0.743 | | 0.114 | | 0.036 | | 0.016 | | 0.003 | | 0.003 | | 0 | | 0 | | 0 | | 0 |
| **20-24** | 0.156 | | 0.369 | | 0.172 | | 0.106 | | 0.033 | | 0.037 | | 0.02 | | 0.016 | | 0.027 | | 0.01 |
| **25-29** | 0.058 | | 0.206 | | 0.218 | | 0.194 | | 0.12 | | 0.065 | | 0.072 | | 0.046 | | 0.038 | | 0.007 |
| **30-34** | 0.033 | | 0.15 | | 0.227 | | 0.26 | | 0.195 | | 0.12 | | 0.077 | | 0.045 | | 0.038 | | 0 |
| **35-39** | 0.005 | | 0.046 | | 0.139 | | 0.193 | | 0.231 | | 0.183 | | 0.092 | | 0.086 | | 0.023 | | 0.212 |
| **40-44** | 0.005 | | 0.049 | | 0.073 | | 0.115 | | 0.177 | | 0.201 | | 0.142 | | 0.152 | | 0.25 | | 0.056 |
| **45-49** | 0 | | 0.026 | | 0.077 | | 0.07 | | 0.085 | | 0.135 | | 0.196 | | 0.217 | | 0.122 | | 0.387 |
| **50-54** | 0 | | 0.013 | | 0.03 | | 0.025 | | 0.048 | | 0.088 | | 0.131 | | 0.155 | | 0.247 | | 0.051 |
| **55-59** | 0 | | 0.021 | | 0.024 | | 0.021 | | 0.012 | | 0.14 | | 0.072 | | 0.24 | | 0.151 | | 0.12 |
| **60-64** | 0 | | 0.006 | | 0.003 | | 0 | | 0.095 | | 0.027 | | 0.197 | | 0.043 | | 0.104 | | 0.156 |
| **Total** | 1 | | 1 | | 1 | | 1 | | 1 | | 1 | | 1 | | 1 | | 1 | | 1 |
| ***Casual partners*** | |  | |  | |  | |  | |  | |  | |  | |  | |  | |
| **15-19** | 0.389 | 0.073 | | 0.029 | | 0.017 | | 0.005 | | 0.001 | | 0.008 | | 0.005 | | 0 | | 0.01 | |
| **20-24** | 0.257 | 0.297 | | 0.146 | | 0.08 | | 0.043 | | 0.033 | | 0.022 | | 0.037 | | 0.028 | | 0.096 | |
| **25-29** | 0.138 | 0.196 | | 0.218 | | 0.149 | | 0.091 | | 0.088 | | 0.079 | | 0.065 | | 0.108 | | 0.015 | |
| **30-34** | 0.097 | 0.13 | | 0.18 | | 0.158 | | 0.124 | | 0.11 | | 0.151 | | 0.153 | | 0.15 | | 0.108 | |
| **35-39** | 0.031 | 0.078 | | 0.123 | | 0.139 | | 0.163 | | 0.131 | | 0.22 | | 0.188 | | 0.281 | | 0.141 | |
| **40-44** | 0.004 | 0.048 | | 0.097 | | 0.1 | | 0.106 | | 0.139 | | 0.115 | | 0.189 | | 0.213 | | 0.214 | |
| **45-49** | 0.034 | 0.025 | | 0.067 | | 0.106 | | 0.138 | | 0.089 | | 0.111 | | 0.093 | | 0.102 | | 0.156 | |
| **50-54** | 0.019 | 0.042 | | 0.053 | | 0.103 | | 0.114 | | 0.141 | | 0.09 | | 0.073 | | 0.038 | | 0.202 | |
| **55-59** | 0 | 0.026 | | 0.076 | | 0.089 | | 0.147 | | 0.139 | | 0.086 | | 0.033 | | 0.038 | | 0.046 | |
| **60-64** | 0.032 | 0.085 | | 0.01 | | 0.06 | | 0.069 | | 0.13 | | 0.118 | | 0.164 | | 0.042 | | 0.012 | |
| **Total** | 1 | 1 | | 1 | | 1 | | 1 | | 1 | | 1 | | 1 | | 1 | | 1 | |

**Table S4.** Model parameters relating to HIV and gonorrhoea.

| **Parameter** | **Value** | **Source** |
| --- | --- | --- |
| Duration acute HIV infection | 1 month | (10) |
| Time from HIV infection until HIV can be detected | 3 months | (17) |
| Duration untreated HIV infection, until AIDS development | 10 years | (17) |
| Viral load during acute HIV | 7 log10 copies/mL | (10) |
| Multiplicative factor increasing the probability of HIV transmission when the HIV-positive man has also gonorrhoea | 1.17 | (8) |
| Duration untreated gonorrhoea | 180 days | Assumption |
| Duration symptomatic gonorrhoea:  Until symptoms develop  Between symptoms onset and seeking test  Between test and cure | 8 days  7 days  2 days** | (18)  Assumption  (18) |
| % symptomatic gonorrhoea getting treated | 100% | Assumption |
| Duration asymptomatic gonorrhoea  From infection until becoming infectious  From opportunistic test till cure | 8 days  21 days** | (18)  Assumption |

*Estimated from data from Stichting HIV Monitoring (SHM).

** Symptomatic NG is immediately treated but not immediately cured; After an opportunistic test, there is some time between the test, test result, and new appointment for treatment.

**Table S5:** Testing distributions of MSM according to known HIV status and number of partners in the preceding 6 months.

| **Testing scenario** |  | **Testing frequency** | **Number of partners in preceding six months** | | | | |
| --- | --- | --- | --- | --- | --- | --- | --- |
|  |  |  | **0-2** | **3-4** | **5-9** | **10-20** | **>20** |
| **Small increase among all MSM** | **HIV-negative & undiagnosed PLWHA** | Every 5 years | 66% | 57% | 50% | 43% | 40% |
|  |  | Every 2 years | 22% | 27% | 32% | 33% | 31% |
|  |  | Every 6 months | 12% | 15% | 18% | 24% | 29% |
|  | **Diagnosed PLWHA** | Every 5 years | 56% | 46% | 35% | 32% | 30% |
|  |  | Every 2 years | 26% | 32% | 36% | 36% | 36% |
|  |  | Every 6 months | 18% | 23% | 29% | 32% | 34% |

*Percentages are among the 80% of MSM getting tested for HIV/STI; the remaining 20% of MSM do not get tested. HIV-negative individuals and undiagnosed persons living with HIV/AIDS (PLWHA) get tested for HIV and gonorrhoea; diagnosed PLWHA get tested only for gonorrhoea. HIV status indicates the known status.

1.	Heijman T, Geskus RB, Davidovich U, Coutinho RA, Prins M, Stolte IG. Less decrease in risk behaviour from pre-HIV to post-HIV seroconversion among MSM in the combination antiretroviral therapy era compared with the pre-combination antiretroviral therapy era. AIDS. 2012;26(4):489-95.

2.	Heymans R, A. Matser A, Bruisten SM, Heijman T, Geskus RB, Speksnijder AG, et al. Distinct Neisseria gonorrhoeae transmission networks among men who have sex with men in Amsterdam, the Netherlands. The Journal of Infectious Diseases. 2012;206(4):596-605.

3.	van Sighem AIW, F.W.N.M.; Boyd A.; Smit, C.; Matser, A.; van der Valk, M. Monitoring Report 2021. Human Immunodeficiency Virus (HIV) Infection in the Netherlands. Amsterdam: Stichting hiv monitoring; 2021.

12.	Van Empelen P, Van Berkel M, Roos E, Zuilhof W. Schorer monitor 2011. Amsterdam: Schorer. 2011.

13.	den Daas C, Zuilhof W, van Bijnen A, Vermey K, Dorfler T, de Wit J. Survey Mannen & seksualiteit 2018.. Seksualiteit en gezondheid-handelen en denken van MSM in Nederland. 2019.
